# Simulating the impact of different vaccination policies on the COVID-19 pandemic in New York City

**DOI:** 10.1101/2021.01.21.21250228

**Authors:** Wan Yang, Sasikiran Kandula, Jeffrey Shaman

## Abstract

**Purpose:** To analyze potential COVID-19 epidemic outcomes in New York City under different SARS-CoV-2 virus circulation scenarios and vaccine rollout policies from early Jan 2021 to end of June 2021.

**Key findings:** In anticipation of the potential arrival and dominance of the more infectious SARS-CoV-2 variant:

1. Mass-vaccination would be critical to mitigating epidemic severity (26-52% reduction in infections, hospitalizations, and deaths, compared to no vaccination, provided the new UK variant supplants currently circulating variants).
2. Prioritizing key risk groups for earlier vaccination would lead to greater reductions in hospitalizations and deaths than infections. Thus, in general this would be a good strategy.
3. Current vaccination prioritization policy is suboptimal. To avert more hospitalizations and deaths, mass-vaccination of all individuals 65 years or older should be done as soon as possible. For groups listed in the same phase, 65+ year-olds should be given first priority ahead of others.
4. Available vaccine doses should be given to the next priority groups as soon as possible without awaiting hesitant up-stream groups.
5. While efficacy of vaccination off-protocol is unknown, provided immune response following a first vaccine dose persists, delaying the 2^nd^ vaccine dose by ∼1 month (i.e. administer the two doses 8 weeks apart) can substantially reduce infections, hospitalizations, and deaths compared to the 3-week apart regimen. Across all scenarios tested here, delaying the 2^nd^ vaccine dose leads to the largest reduction in severe epidemic outcomes (e.g. hospitalizations and deaths). Therefore, to protect as many people as possible, this strategy should be considered if rapid increases in infections, hospitalization or deaths and/or shortages in vaccines were to occur.

## Background

New York City (NYC) was the first COVID-19 epidemic center in the US. During March – May 2020, NYC recorded over 200,000 confirmed COVID-19 cases and over 20,000 COVID-related deaths. Our modeling work^1^ estimated that around 17% of New Yorkers had been infected by the SARS-CoV-2 virus by the end of May 2020, before a series of public health interventions brought the first pandemic wave under control. Starting the week of 6/7/20, NYC began its phased reopening of industries and since then have restricted all industries including schools to a maximal operating capacity of 50% or lower. In addition, face covering has been mandated in public since the week of 4/12/20. These long-term public health interventions and compliance of the public have allowed the city to remain partially open despite the resurgence of COVID-19 starting fall/winter 2020.

In Dec 2020, SARS-CoV-2 vaccines became available and a phased rollout with detailed priority restrictions started on 12/14/20 in NYC. In this report, we summarize model testing of different vaccine rollout scenarios on future COVID-19 epidemic outcomes in the city. These include:

1. Vaccination regimen: 2 doses, 3 weeks apart vs. 2 doses, 8 weeks apart;
2. Pre-testing/infection history interview prior to vaccination: no pre-testing vs. pre-testing that identifies and defers 50% of those with prior immunity;
3. Vaccine uptake: high uptake with 90% among key priority groups to 70% among the general public, median uptake with 70% among key priority groups to 50% among the general public, vs. low uptake with 50% among key priority groups to 30% among the general public; and
4. Alternative vaccination prioritization policy: current policy per CDC guidelines that places 65-74 year-olds in Phase 1c (see Table 1 below for detail) vs. alternative that moves up 65-74 year-olds to Phase 1b, immediately after 75+ year-olds.

**Table 1.** Vaccination prioritization policy in NYC per CDC guideline, as of 1/12/21.

| Phase | Eligible Groups |
| --- | --- |
| 1a | Health care workers; Residents and Staff in Certain Group Living Facilities |
| 1b | 75+ year-olds; Grocery workers; First Responders and Support Staff for First Responder Agencies; Corrections; P-12 Schools, College and Child Care; Public Transit; Homeless Shelters; Other frontline essential workers; Other at-risk groups |
| 1c | 65-74 year-olds; People with certain underlying health conditions (to be determined by New York State)<br>All other essential workers (to be determined by New York State) |
| 2 | All other people |

In addition, in mid-December 2020, a new SARS-CoV-2 virus variant was reported in the UK and quickly became dominant there and has been reported in other countries. Multiple lines of evidence suggest that this new variant is more infectious.^2, 3^ These include 1) cohort matching analysis showing ∼50% higher secondary attack rate among contacts with those infected by the new variant; 2) viral testing and sequencing showing that the new variant became dominant within 1-2 months; 3) higher viral load among those infected with the new variant; and 4) modeling showing that epidemic growth rates in places where the new variant became dominant were ∼40-70% higher than elsewhere. In light of the emergence of this new variant, we model potential epidemic outcomes under the above vaccine rollout scenarios in NYC for two main virus circulation scenarios, separately: 1) the current SARS-CoV-2 variant in NYC remaining dominant in the coming months (as of 1/12/21, the new variant has not been detected in NYC); and 2) the new variant becoming dominant over the course of 4 weeks and remaining so afterwards.

## Methods and main model assumptions

### 1. Model-inference and projection system

The network model-inference system we developed to model SARS-CoV-2 transmission in NYC during the first pandemic wave (i.e. 3/1/20 – 6/6/20) has been described in Yang et al.^1, 4^ In this study, we use an age-specific network SEIRSV (susceptible-exposed-infectious-recovered-susceptible-vacination) model, similar to the model described in Yang et al.^1^ However, here we further include vaccination and waning immunity. Specifically, for the additional vaccination module, we model two doses of vaccine, according to currently available vaccines like the Pfizer-BioNTech vaccine. Based on Polack et al.,^5^ we assume that vaccine efficacy is 85% fourteen days after the first dose and 95% seven days after the second dose. Note that here we use a vaccine efficacy estimate slightly lower than that reported for the first dose (∼90% within 3-4 week of follow-up for the Pfirzer-BioNTech or Moderna vaccines) to account for the potential of lower efficacy outside the 3-4 week window should the 2^nd^ dose be delayed. For the additional waning immunity module, we include a parameter estimating the duration of immunity (with a prior mean of 3 years); see details in Yang et al.^6^ and the Appendix of Yang et al.^1^

In this study, we run the age-specific network SEIRSV model in conjunction with the ensemble adjustment Kalman filter (EAKF)^7^ to calibrate the model system using confirmed and probable COVID-19 case and mortality data from the week of 3/1/20 to the week of 12/27/20 (i.e., the last week with available data at the time of this study). From the week of 1/3/21 to the week of 6/27/21 (i.e. end of June 2021), the model is integrated stochastically using the last estimates of state variables and model parameters. In effect, this setting assumes that there are no changes in public health interventions or behavioral changes during our projection period. However, the number of vaccines administered during the projection period are accounted for according to different rollout scenarios. In addition, the transmission rate and infectious period of SARS-CoV-2 are adjusted for model scenarios assuming introduction and replacement with the new variant (see details below).

We include 8 age-groups (<1, 1-4, 5-14, 15-24, 25-44, 45-64, 65-74, and 75+ year-olds) and all 42 United Hospital Fund (UHF) neighborhoods in NYC. Each simulation (500 ensemble members) is repeated 5 times and aggregated for the summary.

### 2. Modeling vaccine rollout and uptake

As of 1/11/21, 216,014 doses of SARS-CoV-2 vaccines have been administered since 12/14/20 and vaccine rollout is expected to scale up in the coming weeks. However, the level of scale-up and availability of vaccines remain unknown. For this study, we assume that vaccination capacity will increase to 300,000 doses per week by the week of 1/23/21 and will stay so afterwards. We then model the daily numbers of vaccine doses that would be given to 1^st^ and 2^nd^ dose vaccinees, separately. We estimate the numbers based on 1) the day-of-week pattern observed during the first 3 weeks of rollout; 2) total vaccination capacity (i.e. 300,000 per week); 3) distribution of 1^st^ and 2^nd^ dose per person with the two doses administered 3 weeks apart (recommended for the Pfizer-BioNTech vaccine; for simplicity we do not consider other vaccines like Moderna with a 4-week apart regimen) or 8 weeks apart as an off-protocol alternative to increase the number of people receiving initial vaccine protection.

In addition, willingness to receive the vaccine among different groups when they become eligible is unknown. To account for this uncertainty, we model three vaccine uptake scenarios: 1) high uptake, ranging from 90% among key priority groups to 70% among the general public; 2) median uptake, ranging from 70% among key priority groups to 50% among the general public (i.e. 20% lower than the high uptake scenario); and 3) low uptake, ranging from 50% to 30% (i.e. 20% lower than the median uptake scenario). We assume that when up-stream priority groups refuse vaccination, the available doses are given to the next priority groups such that the weekly total is 300,000 for all weeks from 1/23/21 onwards. Figure 1 shows the modeled daily vaccine numbers under different scenarios.

**Fig 1.**
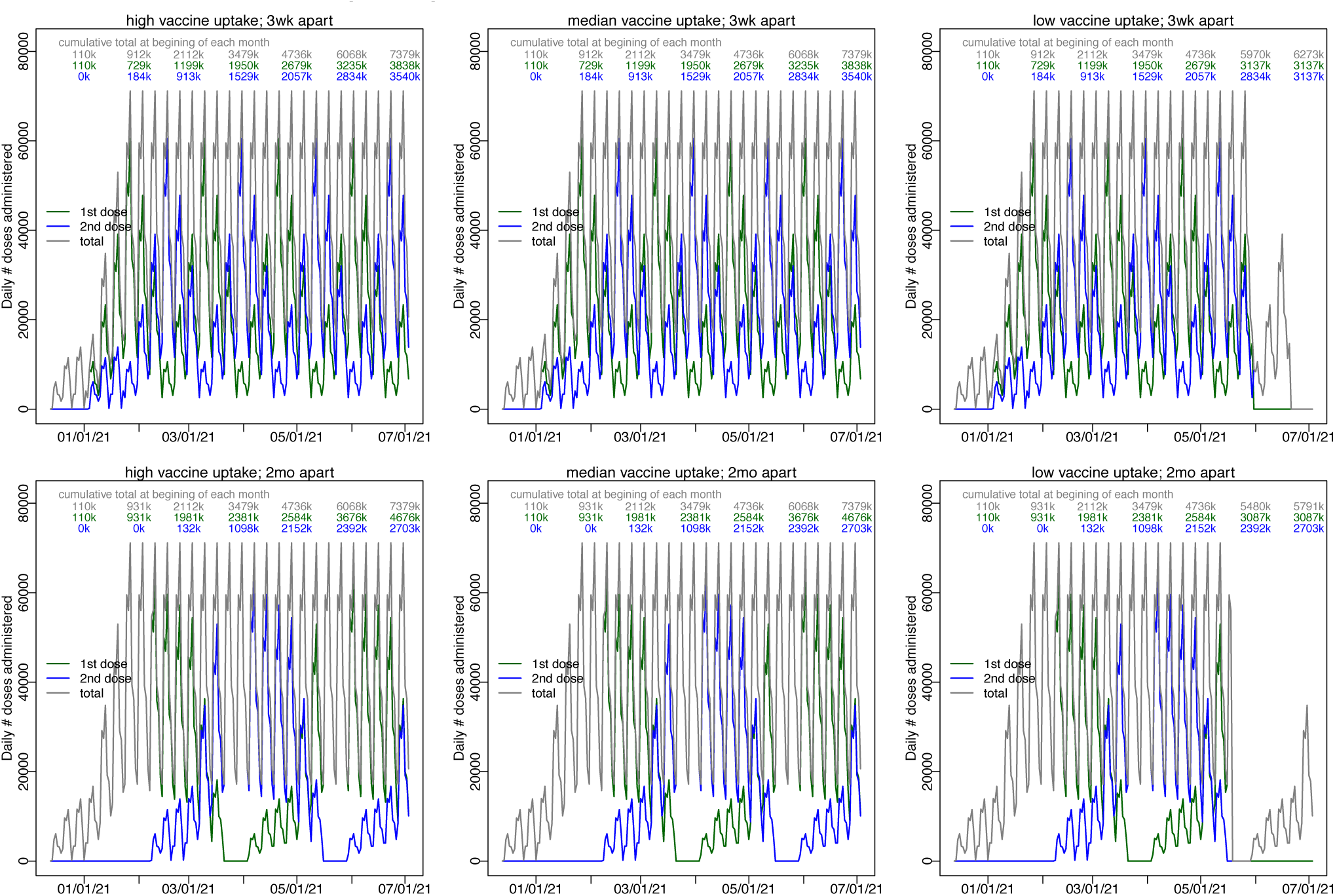
Model estimated vaccination capacity. Total vaccination capacity is kept at 300,000 doses per week starting the week of 1/23/21 (grey lines). Daily numbers of vaccine doses given to 1^st^ dose vaccinees (green lines) and 2^nd^ dose vaccinees (blue lines) are then estimated based on 1) the day-of-week pattern observed during the first 3 weeks of rollout, as well as distribution of 1^st^ and 2^nd^ dose per vaccination regimen either with the two doses administered 3 weeks apart per protocol (top row) or 8 weeks apart as an off-protocol alternative (bottom row). Three vaccine uptake scenarios are modeled: high uptake ranging from 70-90% (1^st^ column), median uptake ranging from 50-70% (2^nd^ column) and low uptake ranging from 30-50% (3^rd^ column). The numbers in the plots show cumulative totals at the beginning of each month.

### 3. Modeling the distributions of vaccinees in different vaccination priority groups

As reported in our previous study,^1^ there are large heterogeneities in the risk of SARS-CoV-2 infection across different age groups and UHF neighborhoods. This leads to heterogeneities in underlying population immunity from prior infections across age groups and space and may affect the actual number of people immunized through vaccination. In addition, the vaccination priority policy is in part based on the risk and severity of infection for different groups. As such, different priority groups would have different underlying immunity status prior to vaccination, in addition to different age and space distributions. And following such a priority-based vaccine rollout, different groups would be differentially protected over time. To account for these heterogeneities and impact on epidemic outcomes, we further model the age (same 8 age groups as in our model described above) and space (same 42 UHF neighborhoods) distribution of each priority group, and distribute the daily vaccine doses based on the priority order (see, e.g., Table 1).

### 4. Other key model settings

1. Replacement by the new SARS-CoV-2 variant. For this scenario, the transmission rate is linearly increased over a 4-week period to 55% higher than the current estimate and the infectious period is linearly increased over the same time period to 10% longer than the current estimate and both remain the same afterwards. Overall, this assumes that by the end of the 4-week replacement period, the reproductive number would increase by 70%.
2. Seasonality: For all simulations, we assume there is a winter time seasonality as estimated in Yang et al. (see Appendix therein).^1^
3. Pre-testing of antibody (or medical interview) to identify and defer vaccination of those with a history of SARS-CoV-2 infection. It is likely that prior infection of SARS-CoV-2 can confer immune protection against subsequent infection for a period of time. Thus, for individuals with recent infection, immediate vaccination may provide little additional protection. As such, delaying vaccination for these individuals may free up available vaccine doses to others with greater need. To test the potential impact of pre-testing and deferring vaccination for individuals with prior immunity, we model two scenarios: i) no pre-testing such that the number of people removed from the susceptible pool after vaccination is scaled by the susceptibility of the corresponding group (e.g., if 40% of the group had residual immunity due to prior infection, we compute the number removed by multiplying the number of vaccinees by 60%). And ii) pre-testing to identify individuals with prior immunity such that 50% of such individuals are identified and deferred. For simplicity, we model this as an increase in the number of individuals removed from the susceptible pool (e.g., for the example in the no test scenario, with 50% identified and deferred, we compute the number removed by multiplying the number of vaccinees by 80%). In reality, the additional vaccine doses can be given to others. Thus, our model here *underestimates* the impact and benefit of pre-testing.
4. Waning immunity: Our projection period extends to the end of June 2021, over 1 year after the start of the COVID-19 pandemic. As such, individuals infected early on during the pandemic may no longer have immune protection at time of vaccination, due to waning immunity. Thus, we compute the residual prior immunity using a logistic function assuming a 3-year immunity period.
5. Vaccination prioritization policy. We model two scenarios: i) same as listed in Table 1; and ii) moving 65-74 year-olds to phase 1b, next to 75+ year-olds. For both scenarios, except for those in phase 1a, <u>our model gives the vaccines to different sub-groups in the same phase</u> *<u>sequentially</u>* <u>per the order listed in Table 1 or as specified in ii for 65+.</u>

### Main modeling findings and implications

1. If the current SARS-CoV-2 variant remains dominant, our model projections show the epidemic would decline in the coming weeks. As a result, the impact of vaccinations would be similar under different policies (Figure 2). There would be a 6-9% reduction in infections, hospitalizations, and deaths due to vaccination over the next 6 months, across all scenarios (see Figure 3, left panel).

**Fig 2.**
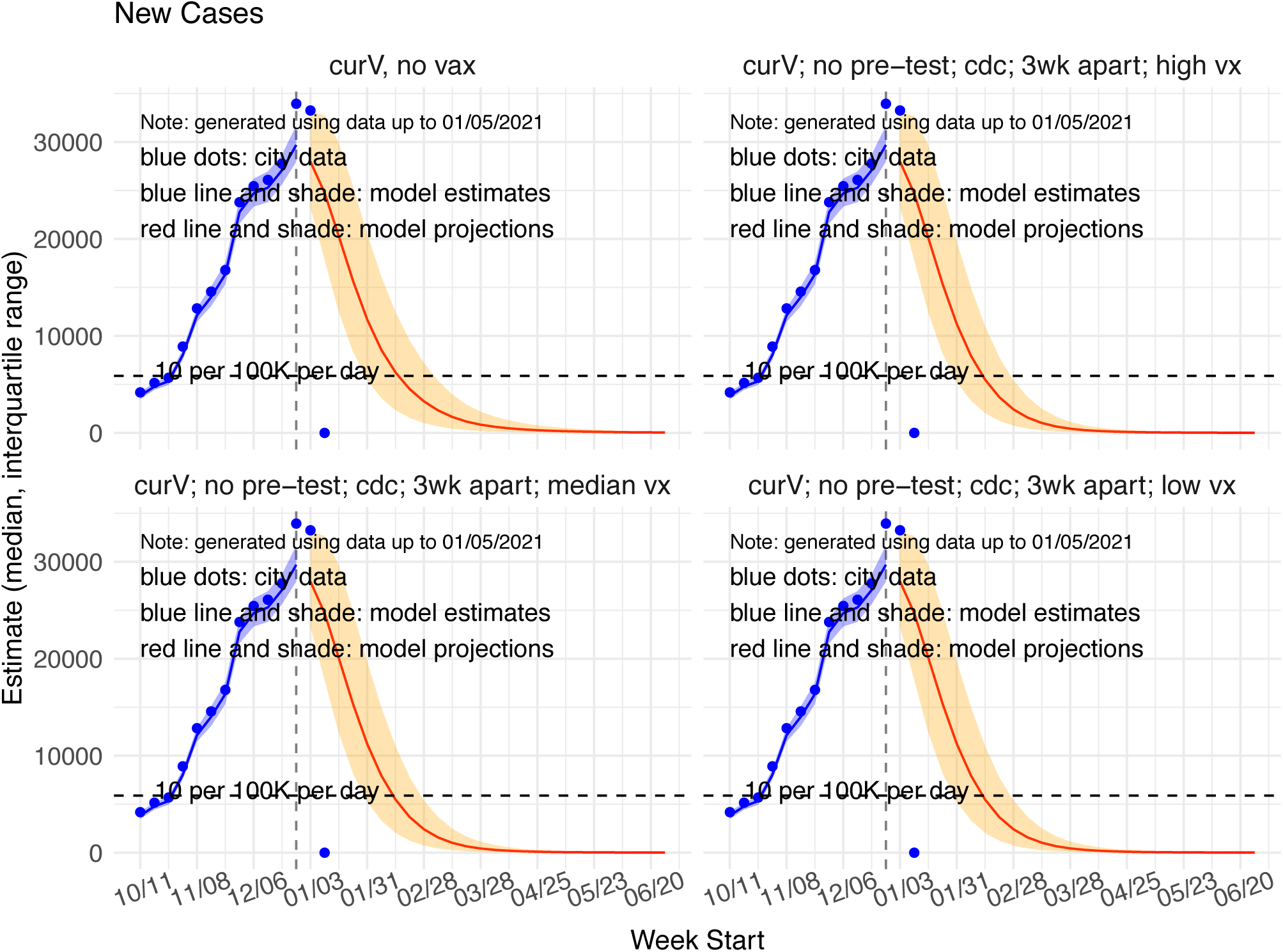
Model estimated and projected weekly numbers of new confirmed and probable COVID-19 cases under the scenario that current SARS-CoV-2 variant remains dominant in the coming months. Blue lines and shaded area show model the estimated median and interquartile range from the week of 10/11/20 to the week of 12/27/20. Red lines and shaded areas show model projections and interquartile ranges from the week of 1/2/21 to 6/27/21. Top left panel shows a baseline scenario with no vaccination. The remaining three panels show vaccine rollout per current policy (i.e. priority groups as listed in Table 1; no pretesting; and 2 doses administered 3 weeks apart) under three vaccine uptake scenarios: high uptake (top right), median uptake (bottom left), and low uptake (bottom right).

**Fig 3.**
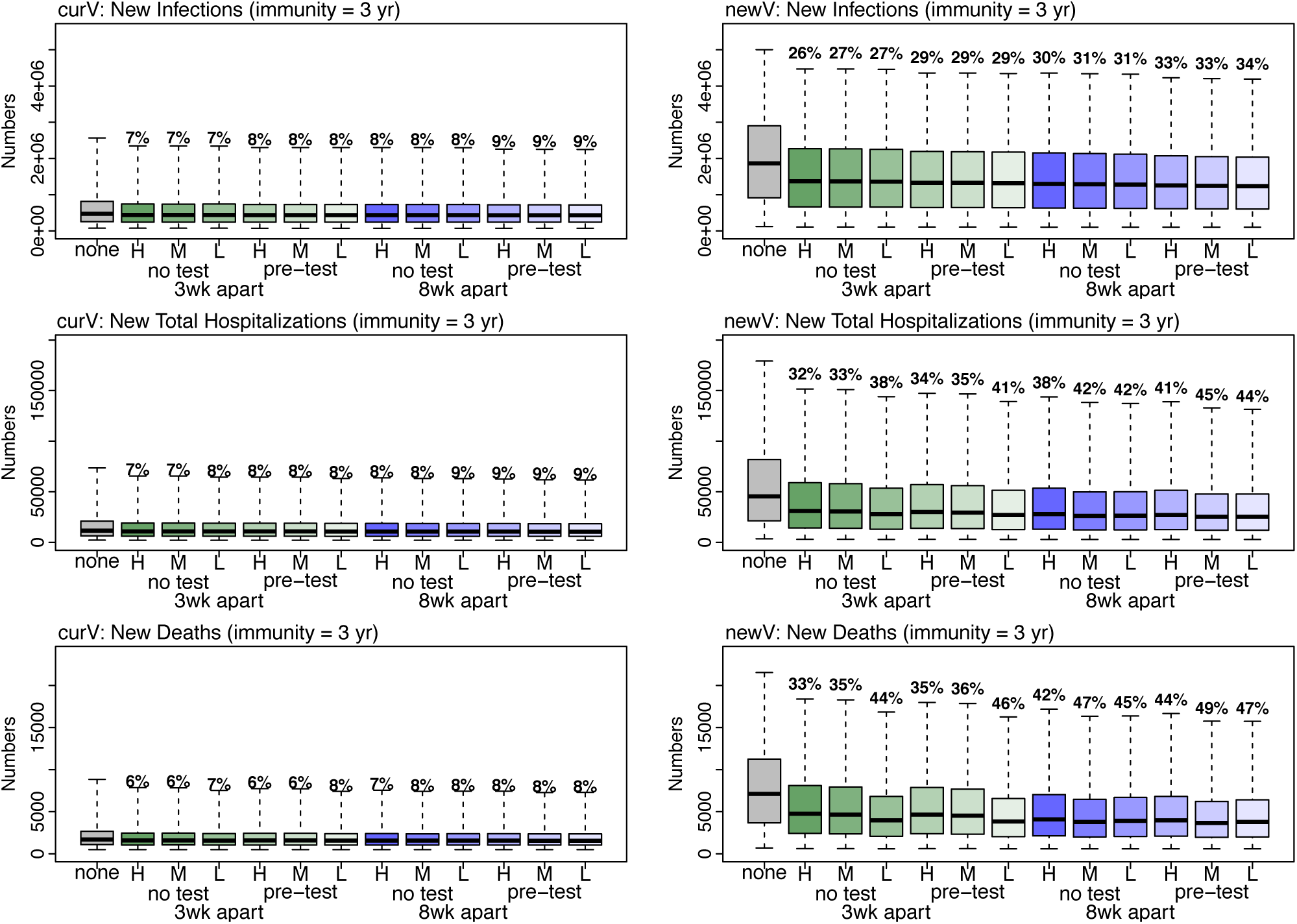
Comparing cumulative epidemic outcomes under different virus circulation and vaccine rollout scenarios. Boxplots show the distribution of the simulated total number of infections (row 1), hospitalizations (row 2), or deaths (row 3), summed over the week of 1/3/21 to the week of 6/27/21 (∼half year). Left panel show model estimates assuming the current SARS-CoV-2 variant remains dominant and right panel show estimates assuming the new variant becomes dominant. Grey boxes show estimates assuming no vaccination; green boxes show estimates assuming 2 vaccine doses administered 3 weeks apart and blue boxes assuming the two doses administered 8 weeks apart. Vaccine uptake scenarios are labeled in the x-axis: H=high uptake; M=median uptake; and L=low uptake; pre-testing scenarios are labeled underneath. Numbers show the estimated median percentage reduction for each health outcome compared to the no vaccination scenario. All scenarios shown here use the vaccination priority per Table 1.
2. If the new variant becomes dominant during the next 4 weeks, substantial increases in infections and other health outcomes would occur (see Figure 3 right panel and Figures 4-6). Without vaccination, the city is likely to have another pandemic wave comparable in magnitude to the 1st pandemic wave in spring 2020. Thus, we will focus on discussing findings assuming the new variant becomes dominant.

**Fig 4.**
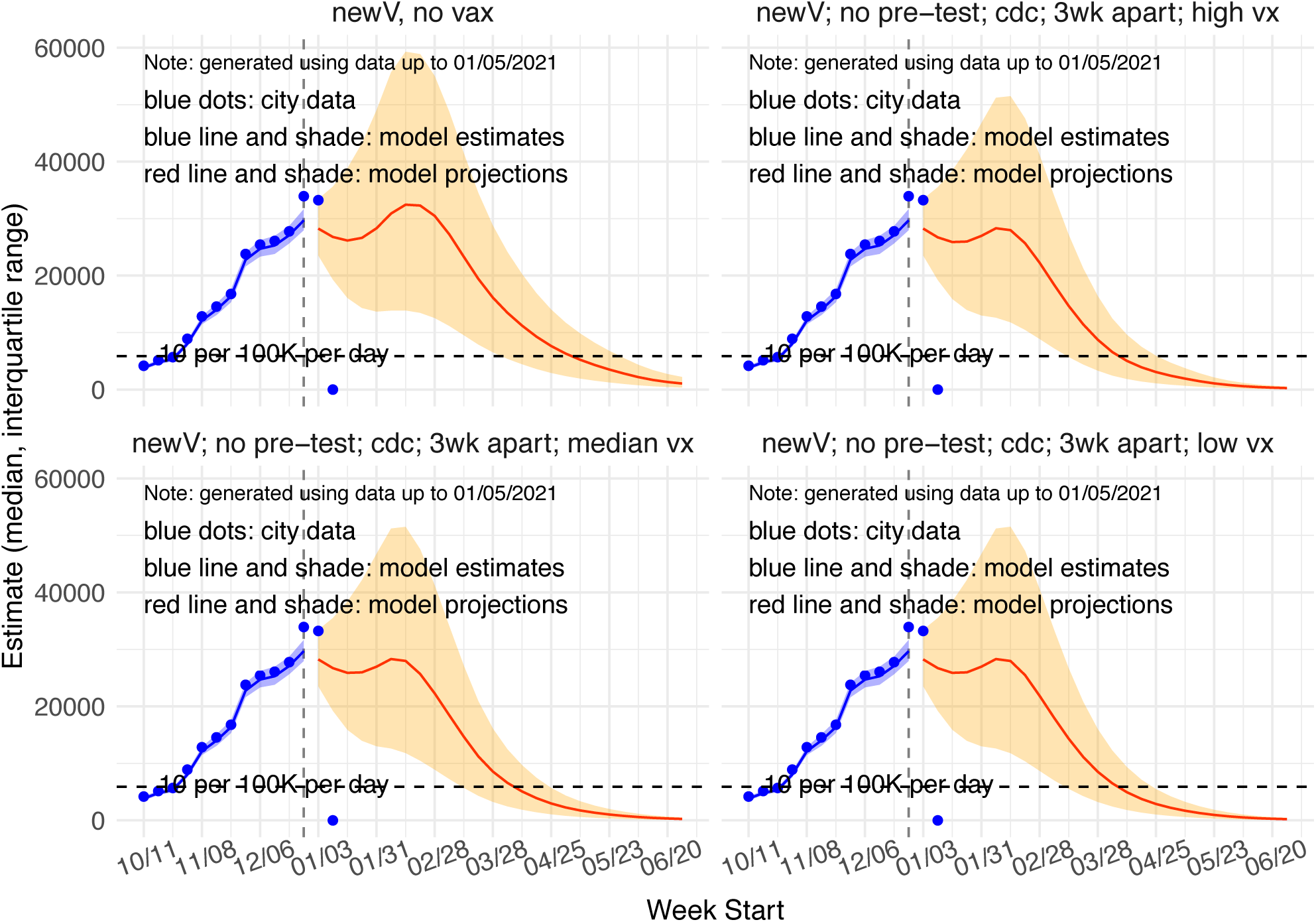
Model estimated and projected weekly number of new confirmed and probable COVID-19 **cases** under the scenario that the new SARS-CoV-2 variant becomes dominant in coming months. Blue lines and shaded areas show model estimated median and interquartile ranges from the week of 10/11/20 to the week of 12/27/20. Red lines and shaded areas show model projections and interquartile ranges from the week of 1/2/21 to 6/27/21. Top left panel shows a baseline scenario with no vaccination. The remaining panels show vaccine rollout per current policy (i.e. priority groups as listed in Table 1; no pretesting; and 2 doses administered 3 weeks apart) under three vaccine uptake scenarios: high uptake (top right), median uptake (bottom left), and low uptake (bottom right). Note that case data were used for model calibration.

**Fig 5.**
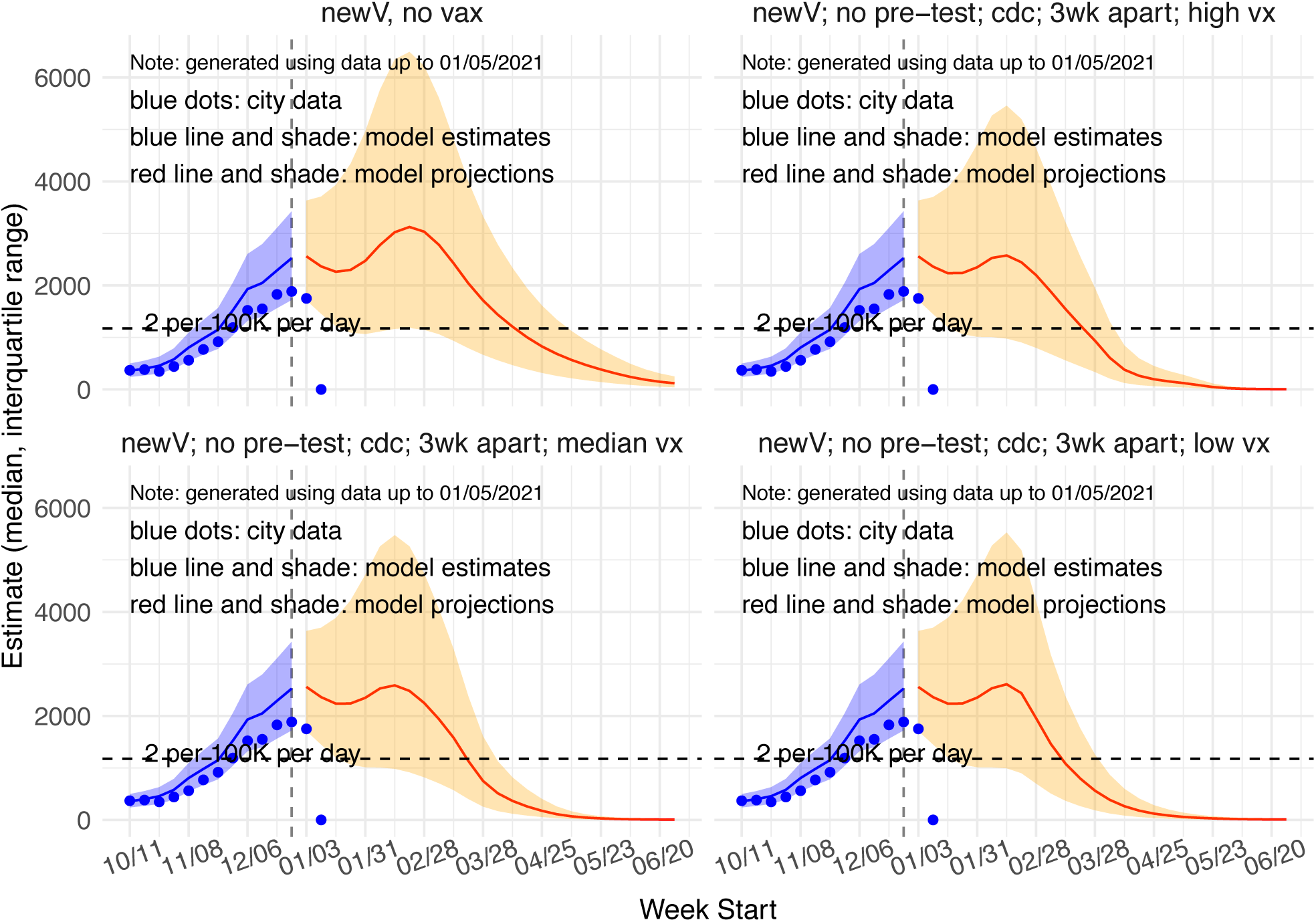
Model estimated and projected weekly number of new confirmed and probable COVID-19 **hospitalizations** under the scenario that the new SARS-CoV-2 variant becomes dominant in coming months. Blue lines and shaded areas show model estimated median and interquartile ranges from the week of 10/11/20 to the week of 12/27/20. Red lines and shaded area show model projections and interquartile ranges from the week of 1/2/21 to 6/27/21. Top left panel shows a baseline scenario with no vaccination. The remaining panels show vaccine rollout per current policy (i.e. priority groups as listed in Table 1; no pretesting; and 2 doses administered 3 weeks apart) under three vaccine uptake scenarios: high uptake (top right), median uptake (bottom left), and low uptake (bottom right). Note that hospitalization data were *not* used for model calibration.

**Fig 6.**
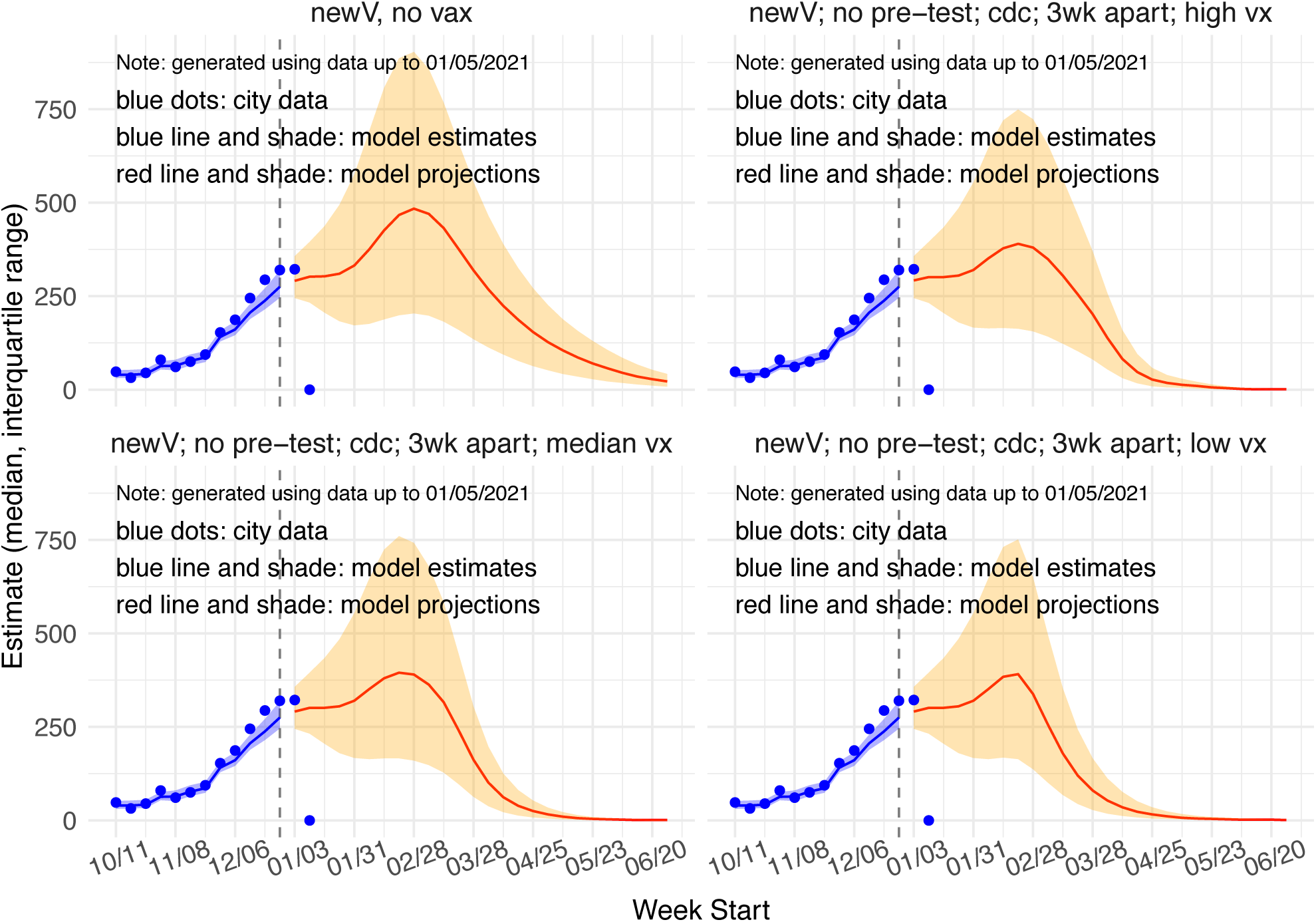
Model estimated and projected weekly number of new confirmed and probable COVID-19 related **deaths** under the scenario that the new SARS-CoV-2 variant becomes dominant in coming months. Blue lines and shaded areas show model estimated median and interquartile ranges from the week of 10/11/20 to the week of 12/27/20. Red lines and shaded areas show model projections and interquartile ranges from the week of 1/2/21 to 6/27/21. Top left panel shows a baseline scenario with no vaccination. The remaining panels show vaccine rollout per current policy (i.e. priority groups as listed in Table 1; no pretesting; and 2 doses administered 3 weeks apart) under three vaccine uptake scenarios: high uptake (top right), median uptake (bottom left), and low uptake (bottom right). Note that mortality data were used for model calibration.
3. With the new variant, vaccination with 300K per week can substantially reduce infections, hospitalizations, and deaths, by 26-52% depending on vaccination strategies and health metrics (see Figure 3, right panel).
4. Prioritizing key risk groups would lead to more substantial reductions in hospitalizations and deaths than in infections (Figure 3, right panel, compare 1^st^ row to 2^nd^ and 3^rd^ rows). Thus, it suggests that in general such prioritization of key risk groups is a good strategy.
5. Current vaccination priority policy is likely suboptimal. Under current vaccination prioritization policy and scenarios with different vaccine uptake rates, the median and low uptake scenarios result in greater reductions than the high uptake scenario. For example, for mortality, there would be a 33% reduction with high uptake vs. a 35% reduction with median uptake and a 44% reduction with low uptake, for the 3-week apart regimen (see Figure 3, bottom-right panel, first 4 boxes). This counterintuitive finding occurs in our model when up-stream priority groups refuse vaccination and the available doses are given to the next eligible priority groups. As a result, this leads to more 65-74 year-olds being vaccinated when uptake is low, as this group has a lower priority than others with occupational risk (as of the time of this study). **These findings suggest that, to avert more hospitalizations and deaths, mass-vaccination of 65+ year-olds should be done as soon as possible. Thus, for groups listed in the same priority phase, 65+ year-olds should be given first priority ahead of others. In addition, available vaccine doses should be given to the next priority groups as soon as possible without awaiting hesitant up-stream groups**.
6. <u>Moving the 65-74 year-olds up the priority list could avert 645 (median, interquartile range: 250 – 1353; same below) more hospitalizations and 129 (61, 284) deaths than current priority policy, assuming median vaccine uptake (see Table 2).</u>

**Table 2.** Impact of prioritizing the elderly 65 or older: Total number of infections, hospitalizations, and deaths projected over 1/3/21 – 7/3/21 under different vaccine uptake scenarios. All simulations here assume no pre-testing. The numbers are the estimated median (and interquartile range). Estimates in the last column (No. averted using the alternative priority policy) are computed as the tally of infections, hospitalizations, or deaths under current vaccination priority policy (as listed in Table 1) minus the corresponding tally under the alternative priority policy moving 65-74 year-olds to phase 1b next to 75+ year-olds.
7. **Delaying the 2**^**nd**^ **vaccine dose by ∼1 month (i**.**e. administering the two doses 8 weeks apart) can substantially reduce infections, hospitalizations, and deaths compared with vaccination 3-weeks apart**. For example, there would be a 47% reduction in deaths under the 8-week apart regimen vs. a 35% reduction in deaths under the 3-week apart regimen, assuming median uptake (see Figure 3, bottom-right panel). <u>Tallies over the 6-month projection period indicate the 8-week apart regimen could avert 4284 (1723 – 8111) more hospitalizations and 872 (376 – 1461) more deaths than current policy, assuming median vaccine uptake (Table 3).</u> Across all scenarios tested here, delaying the 2^nd^ vaccine dose leads to the largest reduction in severe epidemic outcomes (e.g. hospitalizations and deaths). Therefore, while there is uncertainty due to the off-protocol use of vaccine, to protect as many people as possible, this strategy should be considered if there are rapid increases in infections, hospitalizations or deaths, or if there is a shortage of vaccines.

| measure | Vaccine uptake rate | No. total: current vaccination priority policy | No. total: alternative prioritizing 65-74 year-olds | No. averted using alternative priority policy |
| --- | --- | --- | --- | --- |
| Infections (%) | none | 22.2 (10.9, 34.5) | 22.2 (10.9, 34.5) | N/A |
| Infections (%) | high | 16.4 (7.9, 27.1) | 16.4 (7.9, 27.1) | 0 (0, 0) |
| Infections (%) | median | 16.3 (7.9, 27) | 16.3 (7.9, 27) | 0 (0, 0) |
| Infections (%) | low | 16.2 (7.8, 26.8) | 16.3 (7.9, 26.9) | -0.1 (-0.1, 0) |
| Hospitalizations | none | 45407 (21236, 81902) | 45407 (21236, 81902) | N/A |
| Hospitalizations | high | 30973 (14243, 58939) | 30780 (14160, 58505) | 193 (83, 434) |
| Hospitalizations | median | 30454 (14000, 58022) | 29809 (13750, 56668) | 645 (250, 1354) |
| Hospitalizations | low | 28004 (13021, 53513) | 27345 (12794, 52047) | 659 (227, 1466) |
| Deaths | none | 7118 (3669, 11260) | 7118 (3669, 11260) | N/A |
| Deaths | high | 4767 (2427, 8110) | 4733 (2413, 8051) | 34 (14, 59) |
| Deaths | median | 4649 (2374, 7930) | 4520 (2324, 7716) | 129 (50, 214) |
| Deaths | low | 3973 (2079, 6818) | 3818 (2018, 6534) | 155 (61, 284) |

**Table 3.** Impact of delaying the 2^nd^ vaccine dose: Total number of infections, hospitalizations, and deaths projected over 1/3/21 – 7/3/21 under different vaccine uptake scenarios. All simulations here assume the vaccination priority policy per Table 1 and no pre-testing. The numbers are the estimated median (and interquartile range). Estimates in the last column (No. averted using the 8-wk apart regimen) are computed as the tally of infections, hospitalizations, or deaths under the 3-wk apart regimen minus the corresponding tally under the 8-wk apart regimen.
8. Pre-testing can contribute to further reductions in infections, hospitalizations and deaths; but the estimated impact is relatively small (∼2% lower than no pre-testing; see Figure 2). However, as noted above, our model is highly simplified for this effect and as such likely underestimates the benefit of pre-testing.
9. Model uncertainty. Here we assume a constant vaccination capacity of 300,000 total doses per week from the end of Jan 2021 onwards. Should there be decreases in the number of people vaccinated due to vaccine shortage and/or medical staff shortage (e.g., during epidemic peak timing), the number of hospitalizations and deaths averted would likely be lower than estimated here. Conversely, should there be further scale up of vaccination to levels higher than assumed here, the number of hospitalizations and deaths averted would likely be higher than estimated here.
10. Potential relevance to other municipalities. Many municipalities have been experiencing rapid epidemic growth during recent weeks and may continue so in the coming weeks. Given the similar epidemic trends projected under scenarios that the new SARS-CoV-2 variant becomes dominant in NYC, our findings here may also apply to municipalities with continuous, rapid epidemic growth.

| measure | Vaccine uptake rate | No. total: 3-wk apart regimen | No. total: 8-wk apart regimen | No. averted using the 8-wk apart regimen |
| --- | --- | --- | --- | --- |
| Infections (%) | none | 22.2 (10.9, 34.5) | 22.2 (10.9, 34.5) | N/A |
| Infections (%) | high | 16.4 (7.9, 27.1) | 15.5 (7.5, 25.7) | 0.9 (0.4, 1.4) |
| Infections (%) | median | 16.3 (7.9, 27) | 15.3 (7.5, 25.4) | 1 (0.4, 1.6) |
| Infections (%) | low | 16.2 (7.8, 26.8) | 15.2 (7.4, 25.2) | 1 (0.4, 1.6) |
| Hospitalizations | none | 45407 (21236, 81902) | 45407 (21236, 81902) | N/A |
| Hospitalizations | high | 30973 (14243, 58939) | 27965 (12953, 53467) | 3008 (1290, 5472) |
| Hospitalizations | median | 30454 (14000, 58022) | 26170 (12277, 49911) | 4284 (1723, 8111) |
| Hospitalizations | low | 28004 (13021, 53513) | 26324 (12359, 50007) | 1680 (662, 3506) |
| Deaths | none | 7118 (3669, 11260) | 7118 (3669, 11260) | N/A |
| Deaths | high | 4767 (2427, 8110) | 4097 (2131, 7038) | 670 (296, 1072) |
| Deaths | median | 4649 (2374, 7930) | 3777 (1998, 6469) | 872 (376, 1461) |
| Deaths | low | 3973 (2079, 6818) | 3926 (2062, 6696) | 47 (17, 122) |

## Data Availability

N/A

## Acknowledgments

This study was supported by the National Institute of Allergy and Infectious Diseases (AI145883), the National Science Foundation Rapid Response Research Program (RAPID; 2027369), and the NYC Department of Health and Mental Hygiene (DOHMH). We thank the NYC DOHMH for data provision and helpful feedback. We also thank Columbia University Mailman School of Public Health for high performance computing and Safe Graph (safegraph.com) for providing the mobility data.

